# Development of City’s Assessment of Mass Casualty Emergency Response and Action Tool

**DOI:** 10.64898/2026.02.09.26345930

**Authors:** Junaid A. Razzak, Craig Tower, Diksha Mishra, Agnes Usoru, Walid Farooqi, Daniel Barnett, Gai Cole, Jessica Yohana Ramirez Mendosa, Lubna Baig, Maciej Polkowski, Mehrose Ahmad, Edbert B. Hsu

## Abstract

**Background:** The accelerating pace of urbanization worldwide has highlighted the improvement of disaster response in cities as a global priority. Yet, there remains a poor understanding of the emergency response to mass casualty incidents (MCI) in these environments. This study aimed to develop a conceptual framework for cities’ responses and potential policy levers.

**Methods:** We conducted a scoping review followed by in-depth interviews (IDIs), focus group discussions (FGDs), and a modified Delphi process to develop the framework for Cities’ Assessment of Mass Casualty Emergency Response and Action (CAMERA).

**Results:** CAMERA framework consists of six essential components of urban emergency response systems: 1) communication, 2) safety and security, 3) human resources, 4) policy and plans, 5) command control and coordination, and 6) care delivery. IDIs and FGDs also provided insight on assessment methodologies for evaluating response capacity. Using these components, we then developed a framework consisting of a diagnostic and management approach that city leadership can undertake in MCI management to ensure effective functioning at various levels of incident response.

**Conclusion:** The CAMERA framework offers novel and simplified guidance to policymakers and other stakeholders in their attempt to improve MCI response systems across cities globally.

## Introduction

Mass casualty incidents (MCIs) are events that often result in a large surge of victims, overwhelming the capacity of existing health system resources^1^. Globally, MCIs are on the rise due to rapid population and industrial growth, poor infrastructure, poverty, social disruption and climate crises^2^. In recent decades, cities and urban areas in low-and middle-income countries (LMICs) worldwide have become particularly vulnerable to MCIs, leading to events such as fires, structural collapses, motor vehicle crashes, violence, and terrorism^3^. Beyond the immediate loss of life and disruption, MCIs often result in long-term physical and psychological impacts among survivors, imparting an unseen further burden to already limited health resources^4^.

Yet, many cities do not have well developed MCI management plans, leading to preventable morbidity and mortality^4^. Furthermore, existing MCI protocols have largely been designed for high-income countries with considerably greater resources in mind, despite MCIs being more prevalent in LMICs^5^. Effective response to urban MCIs requires highly coordinated efforts by health authorities at all levels— far more complex than standard responses to individual emergencies^6^. The infrastructure to carry out the types and scale of responses needed is often not readily available in LMICs^5^. For example, many LMICs have limited or no organized emergency medical services (EMS); little standardized communication systems between first responders and hospitals and; overcrowded hospitals with limited capacity to manage even modest patient surges due to shortages of personnel, supplies, and MCI training^5^.

Globally, there has been growing interest in strengthening MCI response^7^–13. WHO^14^ and the CDC^15^ have published guidance for policy makers and communities with the aim of identifying system level adaptations needs and interventions. Identifying the key components of urban emergency health response systems to be augmented along with reliable tools to recognize system gaps can help LMIC urban administrators and policymakers improve MCI response. Increasing the readiness of urban emergency health systems for disasters requires addressing four key questions: 1) *What are the essential components of an urban emergency health system?*; 2) *How can gaps among these essential components be identified?*; 3) *Can these gaps be addressed by known field best practices?*; and 4) *Can these findings be formulated to help policymakers systematically assess their city’s disaster readiness, compare performance with benchmarks, and guide future preparedness efforts?* To our knowledge, no universally validated tools exist to identify gaps in the *overall* emergency response capacity of LMIC cities. Accordingly, an assessment tool could offer valuable insights on urban LMIC emergency response capacity.

This study created a conceptual framework to identify critical components for MCI response and develop a tool for measuring urban preparedness. This is much needed to capture the attention of global policymakers, and is particularly salient to achieving sustainable, safe, and resilient cities and communities.

## Methods

This study was led by the Johns Hopkins University (JHU) School of Medicine in collaboration with the International Committee of the Red Cross (ICRC). A framework for mass casualty response was developed from mixed qualitative and quantitative data processes, which included a literature review, in-depth interviews (IDIs), focus group discussions (FGDs), and a modified Delphi process. The approach adopted is well-defined and previously utilized in other studies for research tool development^16,17^.

### Literature Review

A scoping literature review was conducted to identify essential components of an urban emergency response system^18^. A structured search of Pubmed, Embase, Global Health, and Scopus was performed, incorporating four key concepts of (1) mass casualty incidents and synonyms; (2) emergency response and all associated activities (3) performance evaluation methods; and (4) urban population. Search parameters included citations in the English language published between the years 2000-2020. Opinion pieces and abstracts without full text were excluded. Our research focus was on man-made, short-course or sudden MCIs; they did not include natural disasters, bioterrorism events, or pandemics. Seven team members independently screened the titles and abstracts of all retrieved studies with each citation screened by at least (two) team members. This was followed by a full-text review with data extraction using a predefined data extraction sheet. Team members performed a quality assessment of the articles as recommened from the AHRQ Methods Guide for Effectiveness and Comparative Effectiveness Reviews^19^.

### In-depth Interviews & Focus group discussions

Recognizing the need to account for the unique challenges faced by LMICs, semi-structured IDIs and FGDs were conducted with key informants in three LMIC cities—Karachi, Pakistan; Port Harcourt, Nigeria; and Fortaleza, Brazil. Key informants were recruited with the help of local ICRCs and comprised individuals essential to emergency response. These meetings took place from October 2018 to January 2020. Ethical approval to conduct these IDIs and FGDs was facilitated by JHU under IRB number IRB00157208.

At the beginning of each interview and discussion, informed verbal consent was obtained. The interviews and discussions that followed were conducted using pre-developed, open-ended, semi-structured questions from an interview guide created by qualitative data analysis experts from Johns Hopkins University on the research team. These interviews were guided, though not limited, by health system building blocks and goals/outcomes of the World Health Organization (WHO) Health Systems Framework. The introductions provided and general semi-structured questions can be found in Appendix Table 1.

**Table 1:**
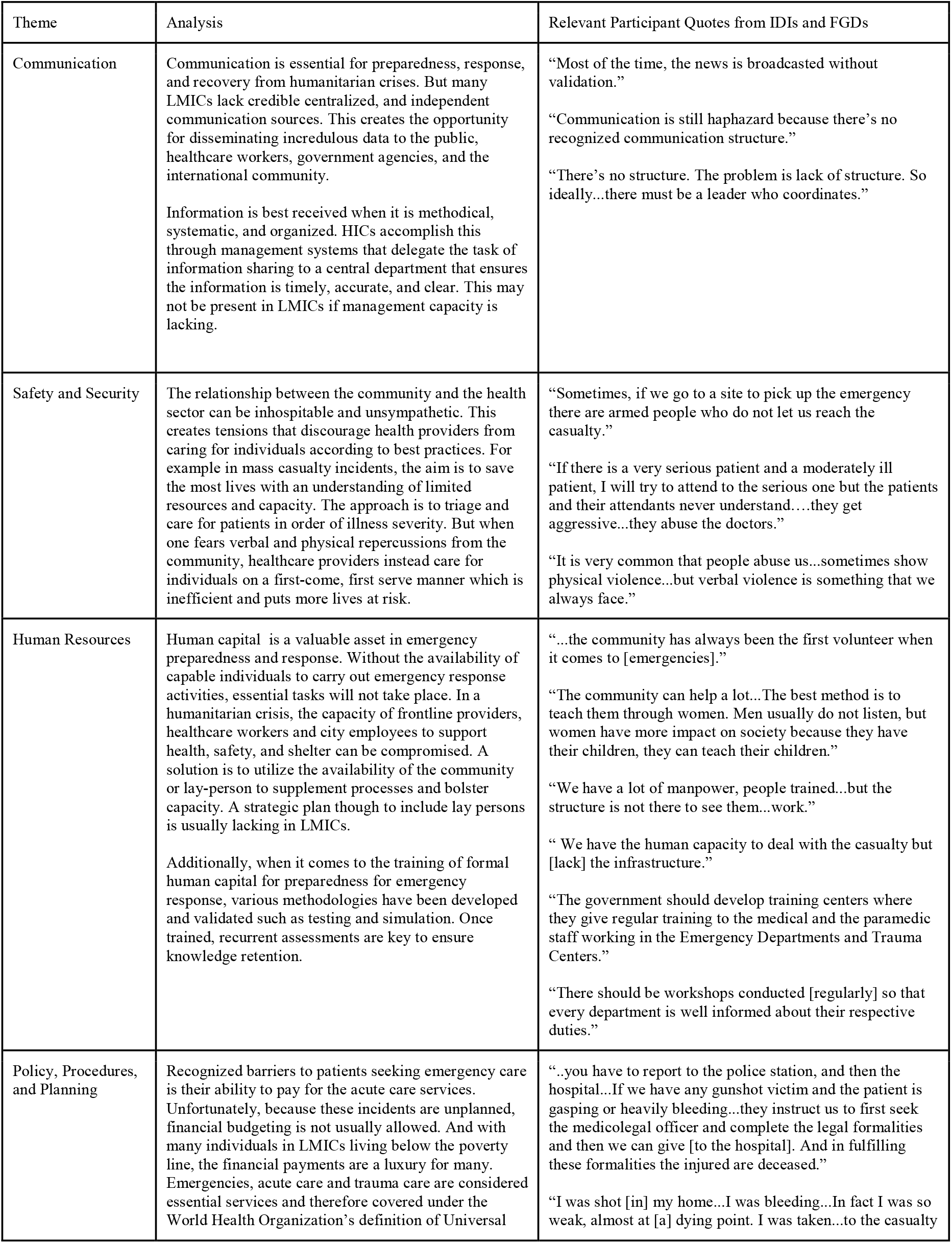
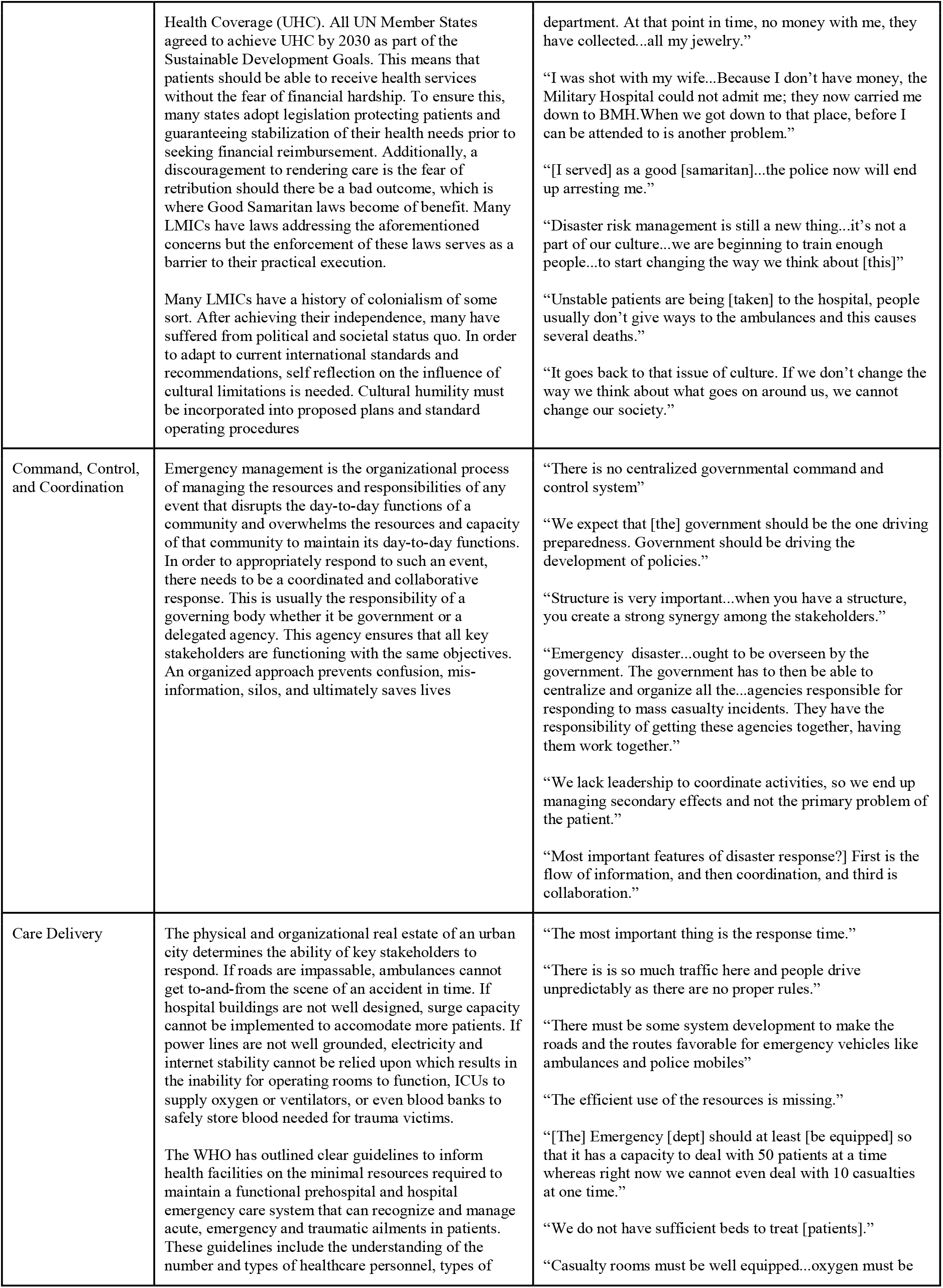
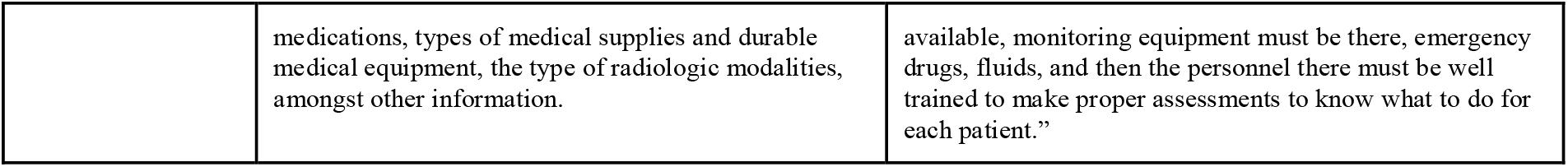
Thematic Analysis of Challenges to LMIC urban emergency response.

| Theme | Analysis | Relevant Participant Quotes from IDIs and FGDs |
| --- | --- | --- |
| Communication | Communication is essential for preparedness, response, and recovery from humanitarian crises. But many LMICs lack credible centralized, and independent communication sources. This creates the opportunity for disseminating incredulous data to the public, healthcare workers, government agencies, and the international community.<br>Information is best received when it is methodical, systematic, and organized. HICs accomplish this through management systems that delegate the task of information sharing to a central department that ensures the information is timely, accurate, and clear. This may not be present in LMICs if management capacity is lacking. | “Most of the time, the news is broadcasted without validation.”<br>“Communication is still haphazard because there’s no recognized communication structure.”<br>“There’s no structure. The problem is lack of structure. So ideally...there must be a leader who coordinates.” |
| Safety and Security | The relationship between the community and the health sector can be inhospitable and unsympathetic. This creates tensions that discourage health providers from caring for individuals according to best practices. For example in mass casualty incidents, the aim is to save the most lives with an understanding of limited resources and capacity. The approach is to triage and care for patients in order of illness severity. But when one fears verbal and physical repercussions from the community, healthcare providers instead care for individuals on a first-come, first serve manner which is inefficient and puts more lives at risk. | “Sometimes, if we go to a site to pick up the emergency there are armed people who do not let us reach the casualty.”<br>“If there is a very serious patient and a moderately ill patient, I will try to attend to the serious one but the patients and their attendants never understand....they get aggressive...they abuse the doctors.”<br>“It is very common that people abuse us...sometimes show physical violence...but verbal violence is something that we always face.” |
| Human Resources | Human capital is a valuable asset in emergency preparedness and response. Without the availability of capable individuals to carry out emergency response activities, essential tasks will not take place. In a humanitarian crisis, the capacity of frontline providers, healthcare workers and city employees to support health, safety, and shelter can be compromised. A solution is to utilize the availability of the community or lay-person to supplement processes and bolster capacity. A strategic plan though to include lay persons is usually lacking in LMICs.<br>Additionally, when it comes to the training of formal human capital for preparedness for emergency response, various methodologies have been developed and validated such as testing and simulation. Once trained, recurrent assessments are key to ensure knowledge retention. | “...the community has always been the first volunteer when it comes to [emergencies].”<br>“The community can help a lot...The best method is to teach them through women. Men usually do not listen, but women have more impact on society because they have their children, they can teach their children.”<br>“We have a lot of manpower, people trained...but the structure is not there to see them...work.”<br>“We have the human capacity to deal with the casualty but [lack] the infrastructure.”<br>“The government should develop training centers where they give regular training to the medical and the paramedic staff working in the Emergency Departments and Trauma Centers.”<br>“There should be workshops conducted [regularly] so that every department is well informed about their respective duties.” |
| Policy, Procedures, and Planning | Recognized barriers to patients seeking emergency care is their ability to pay for the acute care services. Unfortunately, because these incidents are unplanned, financial budgeting is not usually allowed. And with many individuals in LMICs living below the poverty line, the financial payments are a luxury for many. Emergencies, acute care and trauma care are considered essential services and therefore covered under the World Health Organization’s definition of Universal | “...you have to report to the police station, and then the hospital...If we have any gunshot victim and the patient is gasping or heavily bleeding...they instruct us to first seek the medicolegal officer and complete the legal formalities and then we can give [to the hospital]. And in fulfilling these formalities the injured are deceased.”<br>“I was shot [in] my home...I was bleeding...In fact I was so weak, almost at [a] dying point. I was taken...to the casualty |
|  | Health Coverage (UHC). All UN Member States agreed to achieve UHC by 2030 as part of the Sustainable Development Goals. This means that patients should be able to receive health services without the fear of financial hardship. To ensure this, many states adopt legislation protecting patients and guaranteeing stabilization of their health needs prior to seeking financial reimbursement. Additionally, a discouragement to rendering care is the fear of retribution should there be a bad outcome, which is where Good Samaritan laws become of benefit. Many LMICs have laws addressing the aforementioned concerns but the enforcement of these laws serves as a barrier to their practical execution.<br>Many LMICs have a history of colonialism of some sort. After achieving their independence, many have suffered from political and societal status quo. In order to adapt to current international standards and recommendations, self reflection on the influence of cultural limitations is needed. Cultural humility must be incorporated into proposed plans and standard operating procedures | department. At that point in time, no money with me, they have collected...all my jewelry.”<br>“I was shot with my wife...Because I don’t have money, the Military Hospital could not admit me; they now carried me down to BMH. When we got down to that place, before I can be attended to is another problem.”<br>“[I served] as a good [samaritan]...the police now will end up arresting me.”<br>“Disaster risk management is still a new thing...it’s not a part of our culture...we are beginning to train enough people...to start changing the way we think about [this]”<br>“Unstable patients are being [taken] to the hospital, people usually don’t give ways to the ambulances and this causes several deaths.”<br>“It goes back to that issue of culture. If we don’t change the way we think about what goes on around us, we cannot change our society.” |
| Command, Control, and Coordination | Emergency management is the organizational process of managing the resources and responsibilities of any event that disrupts the day-to-day functions of a community and overwhelms the resources and capacity of that community to maintain its day-to-day functions. In order to appropriately respond to such an event, there needs to be a coordinated and collaborative response. This is usually the responsibility of a governing body whether it be government or a delegated agency. This agency ensures that all key stakeholders are functioning with the same objectives. An organized approach prevents confusion, mis-information, silos, and ultimately saves lives | “There is no centralized governmental command and control system”<br>“We expect that [the] government should be the one driving preparedness. Government should be driving the development of policies.”<br>“Structure is very important...when you have a structure, you create a strong synergy among the stakeholders.”<br>“Emergency disaster...ought to be overseen by the government. The government has to then be able to centralize and organize all the...agencies responsible for responding to mass casualty incidents. They have the responsibility of getting these agencies together, having them work together.”<br>“We lack leadership to coordinate activities, so we end up managing secondary effects and not the primary problem of the patient.”<br>“Most important features of disaster response?] First is the flow of information, and then coordination, and third is collaboration.” |
| Care Delivery | The physical and organizational real estate of an urban city determines the ability of key stakeholders to respond. If roads are impassable, ambulances cannot get to-and-from the scene of an accident in time. If hospital buildings are not well designed, surge capacity cannot be implemented to accommodate more patients. If power lines are not well grounded, electricity and internet stability cannot be relied upon which results in the inability for operating rooms to function, ICUs to supply oxygen or ventilators, or even blood banks to safely store blood needed for trauma victims.<br>The WHO has outlined clear guidelines to inform health facilities on the minimal resources required to maintain a functional prehospital and hospital emergency care system that can recognize and manage acute, emergency and traumatic ailments in patients. These guidelines include the understanding of the number and types of healthcare personnel, types of | “The most important thing is the response time.”<br>“There is so much traffic here and people drive unpredictably as there are no proper rules.”<br>“There must be some system development to make the roads and the routes favorable for emergency vehicles like ambulances and police mobiles”<br>“The efficient use of the resources is missing.”<br>“[The] Emergency [dept] should at least [be equipped] so that it has a capacity to deal with 50 patients at a time whereas right now we cannot even deal with 10 casualties at one time.”<br>“We do not have sufficient beds to treat [patients].”<br>“Casualty rooms must be well equipped...oxygen must be |
|  | medications, types of medical supplies and durable medical equipment, the type of radiologic modalities, amongst other information. | available, monitoring equipment must be there, emergency drugs, fluids, and then the personnel there must be well trained to make proper assessments to know what to do for each patient.” |

IDIs and FGDs were conducted in English, and audio-recorded and transcribed verbatim by a member of the local research team at each research site into Microsoft Word. These files were then sent electronically to JHU for data analysis using NVivo (version 12, QRS International Pty Ltd). Two independent research team members utilized predefined codes obtained from the literature review analysis to analyze the narrative transcripts until thematic saturation was achieved. Key concepts from qualitative thematic analysis allowed the research team to better understand the distinctive needs, health system context, and policies related to urban emergency response in low-resource settings; in combination with the literature review findings, these themes isolated from IDIs and FGDs promoted the conceptualization of a framework.

### Modified Delphi

A modified Delphi method, completed in January 2020, was utilized to sort the relevant essential components of an urban emergency response system by level of importance through two cycles of feedback rounds of expert review. Each round of review was performed by content experts with expertise and experience in emergency health response using Qualtrics XM software (Qualtrics, Provo, UT).

Advisors from Johns Hopkins University, the ICRC, and the three local sites were asked to nominate experts for inclusion in the Delphi process. Half of all the experts in each round represented LMICs. All themes drawn from the previously described literature review, IDIs, and FGDs were evaluated by the group. The themes were categorized according to response interventions occurring either at the scene, at the hospital, or related to command, control, and coordination, the latter of which usually falls within the responsibility of a designated emergency control center or agency. Round 1 included ranking the themes and reviewing them using the Qualtrics survey. Average scores from round 1 were reviewed and then re-ranked during round 2. Average scores from round 2 were reviewed and re-ranked during round 3 until consensus was obtained.

## Results

### Literature Review, IDIs, FGDs

From 20,456 screened citations, 181 articles met inclusion criteria for analysis. A total of 24 stakeholders participated in IDIs; 97 stakeholders participated in FGDs. From the literature review, IDIs, and FGDs, thematic analysis identified six essential components of an urban emergency response system, along with assessment methodologies for evaluating response capacity. These six components were communication, safety and security, human resources, policy, procedures and plan, command control and coordination, and care delivery; 40 sub-themes were also found among these components. For example, the theme of human resources comprised all personnel involved in patient management—first responders, physicians, nurses, etc—and their respective training and willingness to participate in an emergency medical response. Similarly, care delivery encompassed sub-themes of search and rescue, triage, response time, essential equipment, life-saving medications and supplies, distribution of casualties, and surge capacity. Communication included communication devices, methods and strategies, and early warning systems for communities regarding risk, such as using social media. Safety and security explored first responder and victim safety in hazardous situations, security protocols during the incident, and possible security risks in violent or terrorist attacks. Policies, procedures, and plans focused on factors that could impact MCI responses by influencing coordination and interaction amongst different agencies, such as resources, information sharing, and MCI response financial and economic burden. The final theme of command, control, and coordination—central to any mass casualty incident (MCI) response—included sub-themes such as central decision-making, resource distribution, information dissemination, and inter-agency coordination. Collectively, the identified themes and sub-themes encompassed a broad and diverse range of components essential to effective MCI responses.

These isolated themes and sub-themes were then used by two team members to conduct a thematic analysis of selected literature articles (Table 1); through this process, the most frequently mentioned and essential components of emergency medicine response in LMICs were identified.

### Modified Delphi Process

A total of 41 experts were initially invited by email to participate in the study. Of those invited, 32 accepted and were sent a link to the initial survey developed with the online survey platform Qualtrics (Provo, UT). A total of 27 participants completed this initial survey round and were then invited to participate in survey round two; 25 participants completed second round surveys. The first round of the study presented participants with factors organized into three categories which were sequentially displayed: scene factors (15); city command and coordination factors (7); and hospital factors (16). At the end of each category, participants were asked to add any comments they had about the factors.

Participants were also asked to rate the importance of each factor in saving lives during a mass casualty response on a scale of 1 to 7. The median score for all factors was 5.59, with the lowest being 4.43, and the highest 6.50. Given lack of variation in scoring, all first-round factors were included in the second round of surveys, along with three additional factors (1 coordination, 2 hospital) that were suggested by participants in round one. Accordingly, the second survey included 15 scene factors, 8 coordination factors, and 18 hospital factors. No comments were elicited in the second survey. In the second round, the median score for all factors was 5.54, with the lowest being 4.16, and the highest 6.72. While greater divergence was noted in the minimum and maximum scores, this variation was not great enough to suggest that any factor was deemed unimportant to saving lives in MCIs, nor was it deemed sufficient to suggest that questions in the final tool should be weighted to account for the importance placed on them by the experts who participated in the Delphi process. The scores and data are presented below in Table 2.

**Table 2:**
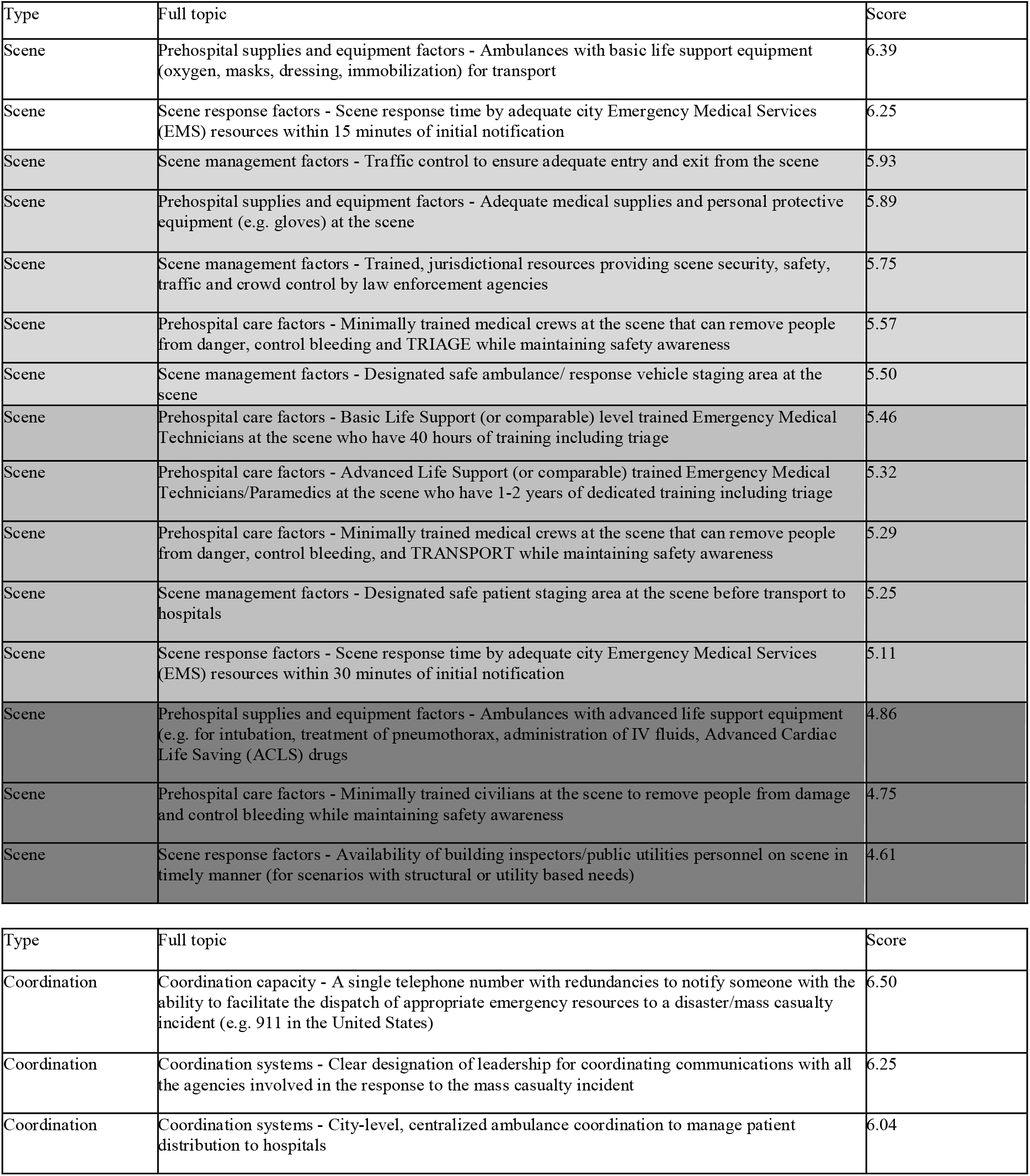
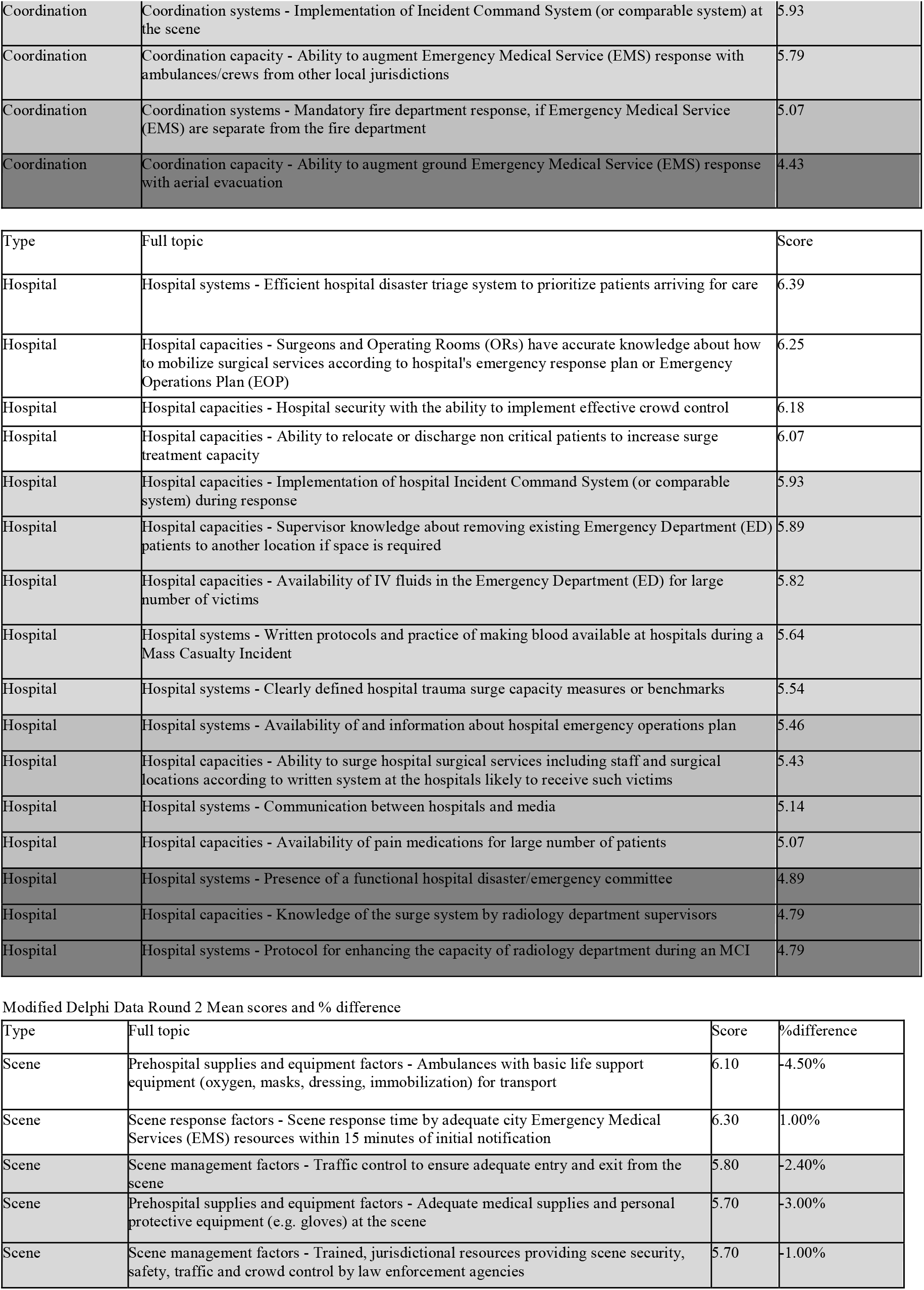
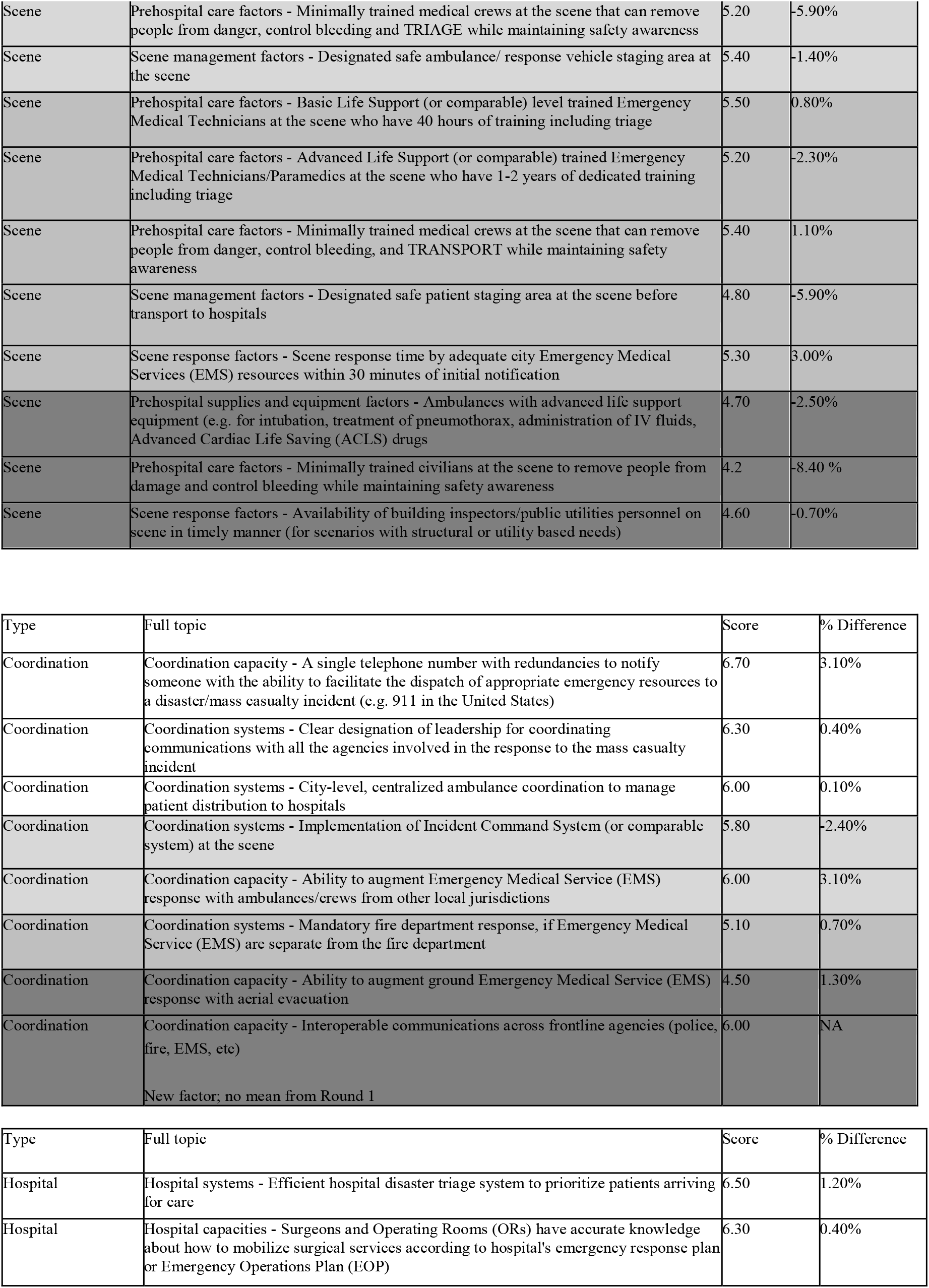
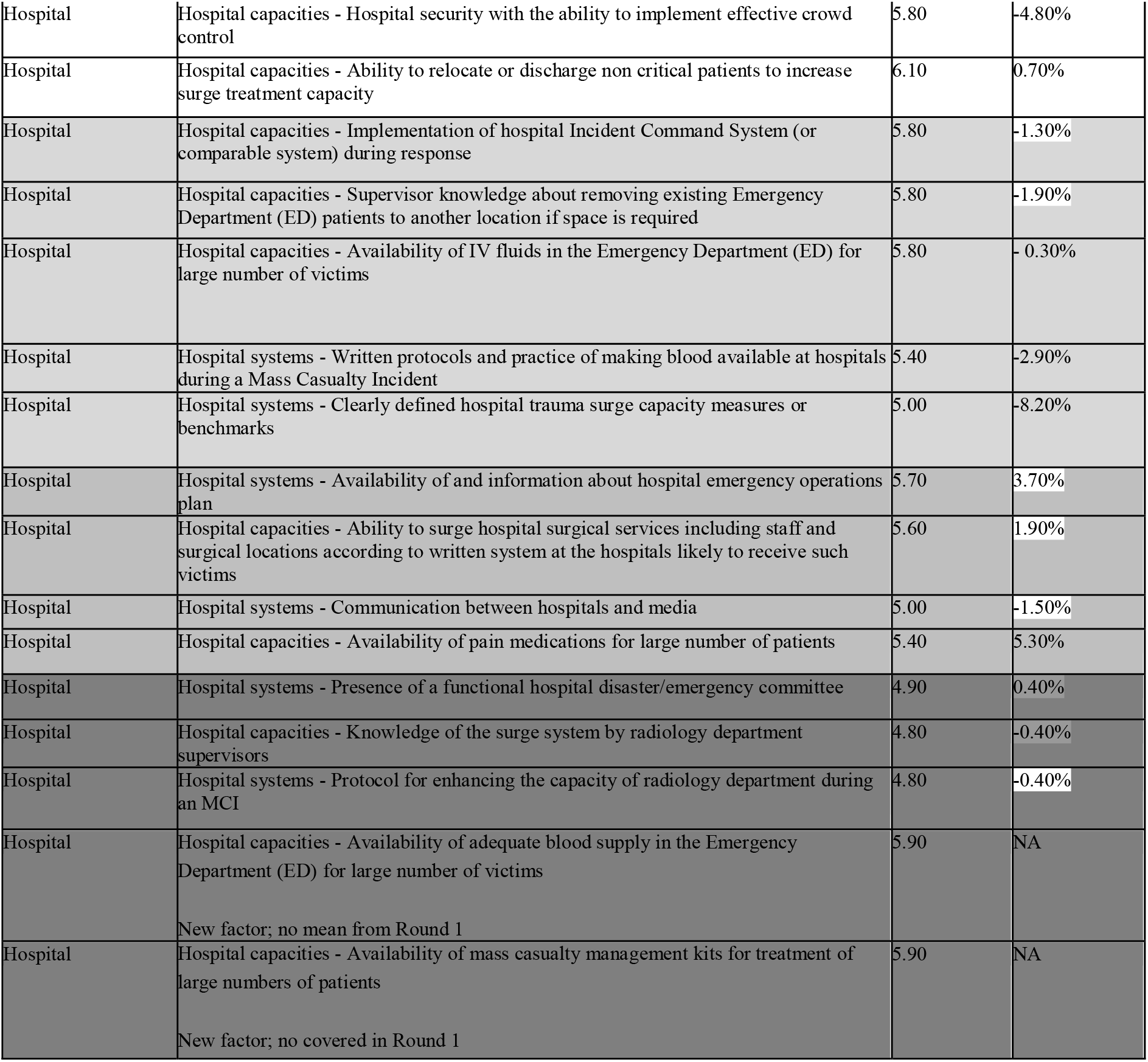
Modified Delphi Data Round 1 Mean Scores.

#### Qualitative Data Analysis

Using our thematic analysis and modified Delphi process as above, we created a framework (Figure 1) that LMIC governments can use to assess their mass casualty response systems. We believe this framework can help policy makers prioritize interventions to address the unique challenges LMICs face in regards to MCI responsiveness, thereby improving response capacity. The red boxes on the left display critical areas at the scene, transport, hospital, and city level that city leadership need to assess their set-up, resources, and protocols in regard to. For instance, initial assessments need to be done to determine if an incident command system (at all levels) is functioning; if there are methods to set up the scene of the disaster effectively, efficiently, and safely; if a means to scale up hospital resources is in place and so forth. All of these aspects have been identified as essential components in responding to an MCI in the literature^11,18,20^–22. By utilizing this diagnostic approach to assess for preparedness, city leadership can then appropriately manage MCIs through formulating and prioritizing a management approach at the level of governance; command control and communication; clinical care, and safety and security. It is important to keep in mind that all of this can only be successfully undertaken in the context of understanding and working within the limitations of LMICs—be that in regard to financial, human, and spatial resources or other aspects. Only within the correct context will city leadership be able to initiate the changes needed to ideally manage a MCI.

**Figure 1:**
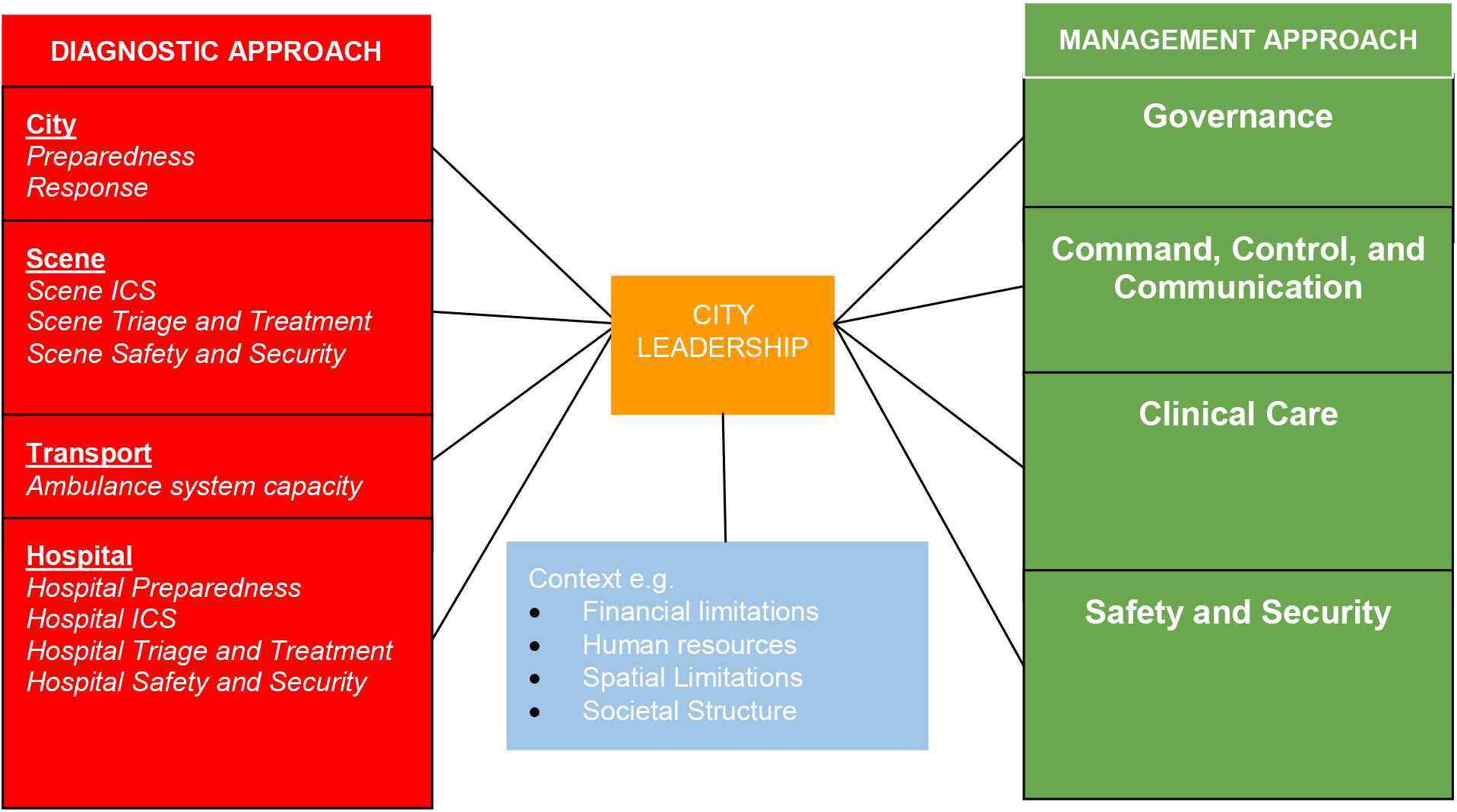
CAMERA Framework for Approach to Mass Casualty Incident Management

## Discussion

By integrating the results from our mixed method data processes and thematic analysis, our team created a framework that displays how to efficiently and effectively approach a mass casualty incident. Our literature review captured a wide-range of research from experimental trials to observation/review articles. Further, our IDIs and FGDs provided the opportunity to learn from experts from LMICs who are directly involved in MCI management, giving current real-world insight into the unique, invaluable needs, challenges, and perspectives that they can offer. Consolidating the themes that emerged from this research and these interactions highlighted six descriptive themes along with numerous subthemes. Rankings of those which were more or less important were derived through the modified Delphi process. All of this was then integrated and incorporated into the framework presented above in Figure 1.

Our framework consists of a diagnostic and management approach to LMIC MCI management, a method that reflects a day-to-day approach to emergency patient care and applies equally well to disaster preparedness overall. City leadership undertakes a diagnostic approach to assess if all the necessary components related to city, scene, transportation, and hospital MCI preparedness and response are present and functioning at an optimal level. Despite the unique challenges that an MCI may bring, each of these areas is vital to being adequately prepared in an LMIC setting. Once all the diagnostic components are accounted for, city leadership can then ensure the appropriate management approach within governance, command control and coordination, clinical care, and safety and security. For instance, governance is related to ensuring that city leadership has all of the policies, protocols, and resources ready to efficiently and effectively handle an MCI. By assuring that all relevant hospital diagnostic needs are met, management of MCIs at the clinical care level can be optimized, including care provided both at the scene of an MCI and at the hospital. By ensuring that components such as an incident command system, a central emergency response agency, an incident commander, and so on are established at the diagnostic level, city leadership can work toward proper command, control, and coordination at the management level. The same can be applied to safety and security. These can all only be achieved by working within a given city’s unique context.

An effective MCI response focuses on saving lives and minimizes chaos, misinformation, and delays in efficient treatment. Encountering shortcomings during the wake of a MCI itself can be disastrous. Likewise, it is challenging to conduct disaster drills and tabletop exercises given the intensive resources and coordination needed for implementation. Development of an urban MCI management capability tool could aid governing bodies in assessing their capability in a focused and specified manner. Through identifying the essential components of a mass casualty response and developing a conceptual framework to visualize the interconnectedness of the essential components, we hope to offer guidance and direct future work by creation of a tool that can evaluate a LMIC’s MCI capabilities and provide concrete steps that the government can take to build upon their emergency response systems.

### Limitations

Our study acknowledges several limitations. For one, the framework does not discuss vulnerability assessments and recovery related to an MCI because it focuses on the acute and immediate response phase of urban emergency response. However, recovery from mass casualty incidents and preventative measures to lessen the impact of future disasters are also essential and necessary for urban resilience. Additionally, our research focus was on man-made, short-course or sudden MCIs that do not compromise the physical integrity of the health system, i.e., natural disasters, and do not jeopardize the human capital necessary to maintain an intact health system, such as a bioterrorism event or a pandemic. The subset of MCIs that we focused upon is increasingly relevant to understanding emergency medical response capacity. Lastly, our analysis is only as strong as the literature available for review. Very few experimental design studies emerged from our search of disaster medicine literature. This is not unexpected due to the random and unpredictable nature of MCIs, which limits opportunities to design and conduct real-time studies. A prominent feature of the current and available disaster medicine research is the focus on process outcomes without correlated health outcomes. Further research on identifying how health outcomes, and other impact outcomes, are affected by improvements in process indicators is needed.

## Conclusion

Using an innovative and mixed-methods approach, we have created a framework that attempts to provide much needed guidance to decision makers as well as other stakeholders to improve mass casualty response systems in cities around the world. This study highlights the components of MCI response that can assist cities in their emergency response efforts. City leadership around the world may feel empowered to better understand and address gaps in their mass casualty response systems. Implementing this framework will set the stage for creation of a tool that can inform governmental bodies of their MCI response capabilities, potentially obviating the need for conducting routine resource-intensive disaster drills.

## Supporting information

Appendix Table 1: Scripts for In-Depth Interviews (IDIs) and Focus Group Discussions (FGDs)

## Data Availability

All data produced in the present study are available upon reasonable request to the authors

