## Appendix Table 1: Scripts for In-Depth Interviews (IDIs) and Focus Group Discussions (FGDs) for "Development of City’s Assessment of Mass Casualty Emergency Response and Action Tool"

**Appendix Table 1: Scripts for In-Depth Interviews (IDIs) and Focus Group Discussions (FGDs)**

| ***Scripted Moderator Introduction for IDIs*** | This interview will contribute to developing a tool using an all-hazards approach to mass casualty events that could occur in the urban environment of low and middle-income countries. A mass casualty event is defined by the World Health Organization as an event that generates more patients at one time than locally available resources can manage using routine procedures. It requires exceptional emergency arrangements and additional or extraordinary assistance.  We need to get your views as experts in emergency response and preparedness. Our goal is to understand what you know about plans in your city to respond to emergencies, what you are supposed to do, and how you communicate with the government, the media, and your community during emergencies. As a result of this discussion, we seek to identify critical determinants of saving lives during the health response to a mass casualty event and ways to improve the chance of survival for victims in your particular city. We will use this information to develop a measurement system. The aim is to help governments of cities like yours worldwide to save lives by improving how they respond to emergencies.  You are being asked to take part in a research study. This study aims to develop and implement an evidence-based assessment strategy for health response systems in urban settings when responding to multiple casualty incidents in low-income settings. If you agree to participate, we will interview you using a semi-structured interview guide, asking questions to identify key themes essential for emergency preparedness and response during a mass casualty incident. The time required to conduct an interview is forty-five minutes to one hour, and the interview will be audiotaped to avoid the inconvenience of missing any useful information. |
| --- | --- |
| ***Scripted Moderator Introduction for FGDs*** | Thank you for agreeing to participate in this discussion. It is important to us to get your views as community leaders. Our goal is to understand what you know about plans in your community to respond to emergencies, what you are supposed to do as a community leader, and how you communicate with the government, the media, and your community during emergencies. As a result of this discussion, we seek to identify critical determinants of saving lives during the health response to a mass casualty event and ways to improve the chance of survival for victims in your particular city. We will use this information to develop a measurement system. The aim is to help governments of cities like yours worldwide to save lives by improving how they respond to emergencies.  You are being asked to take part in a research study. This research study aims to develop and implement an evidence-based assessment strategy for health response systems in urban settings when responding to multiple casualty incidents in low-income settings. If you agree to participate, you will be part of a focus group discussion where we discuss key themes essential for emergency preparedness and response during a mass casualty incident. The time required to conduct this focus group discussion is one hour to one and a half hours, and this activity will be audiotaped to avoid the inconvenience of missing any useful information. |
| ***Scripted Questions for IDIs & FGDs*** | 1. What are the strengths/weaknesses of your current emergency response system? 2. How may you overcome the weakness(es)? 3. What are the most important activities and resources of the government/NGOs/   community/ambulances/prehospital care/law enforcement/hospital/media that would save lives during the first few hours of a mass casualty event?   4. How do you evaluate these? 5. Is there a central group of individuals responsible for decision-making and provision of resources in the first few hours of a mass casualty event? 6. Have you received any disaster-specific training to prepare you to be a better responder during a mass casualty event? If yes, what kind of training did you receive? 7. In the scenario presented in our study case, are there other people from your institution who would be able to know what to do at that moment? 8. Who is responsible in your city for responding to a mass casualty event? 9. Which components of an emergency health response system work better than others? 10. How would you describe the process for maintaining and updating the command and control structure? 11. Can you describe the emergency health response within the first six to ten hours of a mass casualty event in your urban area? How can the response plan be accessed? 12. How are community members in your city included in a mass casualty management process? 13. Is any data on emergency health responses routinely collected? 14. Are there any components of your area's emergency health response system lacking information that would help prepare for a mass casualty event? 15. What procedures are in place to monitor the availability of equipment and supplies before a mass casualty event? 16. How are medical staffing levels tracked? 17. Which stakeholder or stakeholders are responsible for gathering and managing data on emergency preparedness? 18. Who are the stakeholders responsible for collecting and managing this emergency preparedness data? 19. What would be the important features of an ideal emergency response system here? |
